# Multi-omic modelling of inflammatory bowel disease with regularized canonical correlation analysis

**DOI:** 10.1101/2020.04.16.20031492

**Authors:** Lluís Revilla, Aida Mayorgas, Ana Maria Corraliza, Maria C. Masamunt, Amira Metwaly, Dirk Haller, Eva Tristán, Anna Carrasco, Maria Esteve, Julian Panés, Elena Ricart, Juan J. Lozano, Azucena Salas

## Abstract

**Background:** Personalized medicine requires finding relationships between variables that influence a patient’s phenotype and predicting an outcome. Sparse generalized canonical correlation analysis identifies relationships between different groups of variables. This method requires establishing a model of the expected interaction between those variables. Describing these interactions is challenging when the relationship is unknown or when there is no pre-established hypothesis.

**Aim:** To develop a method to find the relationships between microbiome and transcriptome data and the relevant clinical variables in a complex disease, such as Crohn’s disease.

**Results:** We present here a method to identify interactions based on canonical correlation analysis. Our main contribution is to show that the model is the most important factor to identify relationships between blocks. Analysis were conducted on three independent datasets: a glioma, Crohn’s disease and a pouchitis data set. We describe how to select the optimum hyperparameters on the glioma dataset. Using such hyperparameters on the Crohn’s disease data set, our analysis revealed the best model for identifying relationships between transcriptome, gut microbiome and clinically relevant variables. With the pouchitis data set our analysis revealed that adding the clinically relevant variables improves the average variance explained by the model.

**Conclusions:** The methodology described herein provides a framework for identifying interactions between sets of (omic) data and clinically relevant variables. Following this method, we found genes and microorganisms that were related to each other independently of the model, while others were specific to the model used. Thus, model selection proved crucial to finding the existing relationships in multi-omics datasets.

## Background

The creation of datasets from different high-throughput sequencing technologies on the same samples provides an opportunity to identify relationships between datasets and improve our understanding of diseases. This approach has been used in several diseases, such as cancer, inflammatory bowel disease (IBD) and pouchitis, among others [1–3].

IBD is comprised of Crohn’s disease (CD) and ulcerative colitis (UC). Around 4.2 million individuals suffer from IBD in Europe and North America combined [4]. The chronic inflammatory response observed suggests an interaction between host genetic factors and the intestinal microbiota. Several studies support the concept that CD arises from an exacerbated immune response against commensal gut microorganisms in genetically predisposed individuals. Nonetheless, the disease might result from imbalanced microbial composition, leading to dysbiosis [5, 6].

Understanding the contribution of the gut microbiota to CD pathogenesis and maintenance of the disease is an ongoing field of research [7–9]. These alterations could be shaped by a genetic predisposition and environmental factors (i.e., bacterial or viral infection, diet, usage of antibiotic, or the socioeconomic status) [10]. Pouchitis is the inflammation of the ileal pouch, an artificial rectum surgically created out of ileal gut tissue in patients who have undergone a colectomy. One possible underlying cause of pouchitis might be the microbiome [11]. However, the cause-effect relation between dysbiosis and intestinal inflammatory disease remains unclear [12–14].

The most common method for analyzing the relationship between microorganisms and the gut mucosa is to sequence both the 16S rRNA gene of the microbiome and the patient’s transcriptome using DNA and human RNA, respectively, both extracted from the same sample. In some cases, patients are followed up for long periods and longitudinal samples can be obtained [15]. Multiple omics have been increasingly used to identify relationships between the intestinal microbiome and gut epithelium using a variety of methods [8, 14, 16, 17].

Both univariate and multivariate methods are used to analyze DNA and RNA data. Some methods find relationships between the human transcriptome and the gut microbiome composition. Correlations, which are multivariate, are the predominant method used to find relationships between two omics datasets [7, 17–19]. A recent study revealed more significant correlations in samples from healthy controls than in patients with IBD, and suggests an “uncoupling” of the microbiota from homeostasis [7]. Although their analysis used correlations, as well as univariate methods, these method do not consider confounders such as age, diet or sample localization in the gut, all of which could lead to false conclusions [20, 21].

Other multivariate methods provide frameworks with an unlimited number of variables involved. These multivariate methods summarize the variability of the datasets and select features in order to obtain loading factors for a new coordinate system. They aim to summarize the largest amount of variability found among the samples’ variables [22]. Multi-block methods are multivariate methods capable of summarizing several variables from the same sample, but corresponding to different technical origins [23–27]. These multi-block methods assume the existence of relationships between variables of the different blocks.

Regularized generalized canonical correlation analysis (RGCCA) is a multi-block method that enables reducing the dimensions of an arbitrary number of blocks for data derived from the same sample [28–30]. RGCCA has already been used in the context of IBD with RNA-seq and 16S rRNA data [16]. However, it was used to select variables related to the inflammation predictors DUOX2 and APOA1. To our knowledge, a concrete description of the relationship between the gut’s mucosal transcriptome and microbiome in CD using RGCCA has not been performed.

In this study, we evaluate the effect of the parameters of RGCCA and we identify a strategy of analysis that better explains a previously published glioma cohort to validate the method [31]. We then applied this method to our CD dataset and to a pouchitis cohort in order to identify interactions between microorganisms and the transcriptome of the gut epithelium [32]. We believe that this approach is crucial to find the various relationships in multi-omics datasets and select the most relevant variables.

## Methods

### Patients and biopsies processing

Samples from the CD cohort included in this study were from patients treated in the Department of Gastroenterology (Hospital Clínic de Barcelona – Spain –) all of whom signed a consent form. A cohort of patients with severe refractory CD that underwent hematopoietic stem cell transplant (HSCT). Colonic and ileal biopsies were obtained at several time points from CD patients undergoing routine colonoscopies. Patients were followed-up for 4 years and samples were collected every six or twelve months after HSCT. Samples were collected from both uninvolved and involved areas. In addition, biopsies were taken from the ileum and colon of 19 non-IBD patients consisting of individuals with no history of IBD who had no significant pathological findings following endoscopic examination for colon cancer surveillance (Hospital Univesitari Mútua de Terrassa – Spain –). At least one biopsy was fresh-frozen at −80ºC for microbial DNA extraction. The remaining biopsies were placed in RNAlater RNA Stabilization Reagent (Qiagen, Hilde, Germany) and stored at −80ºC until total RNA extraction.

### Mucosal transcriptome

Total RNA from mucosal samples was isolated using the RNeasy kit (Qiagen, Hilde, Germany). RNA sequencing was performed as previously described [15]. Analysis was performed using an R version (3.6.1) statistical tool and Bioconductor (Version 3.9) on Ubuntu 18.04. The transcriptome was visually inspected for batch effects in PCA. Outliers and the top 10% genes using the coefficient of variation were removed. Data was normalized using the trimmed mean of M-values and log transformed into counts per millions using edgeR (version 3.26).

### Microbial DNA extraction from mucosal samples

Biopsies were resuspended in 180 µl TET (TrisHCl 0.02M, EDTA 0.002M, Triton 1X) buffer and 20mg/ml lysozyme (Carl Roth, Quimivita, S.A.). Samples were incubated for 1h at 37ºC and vortexed with 25 µl Proteinase K before incubating at 56ºC for 3h. Buffer B3 (NucleoSpin Tissue Kit – Macherey-Nagel) was added followed by a heat treatment for 10 min at 70°C. After adding 100% ethanol, samples were centrifuged at 11000 × g for 1 min. Two washing steps were performed before eluting DNA. Concentrations and purity were checked using NanoDrop One (Thermo Fisher Scientific). Samples were immediately used or placed at −20°C for long-term storage.

### High throughput 16S ribosomal RNA (rRNA) gene sequencing

Library preparation and sequencing were performed at the Technische Universität München as described in detail previously [33]. Briefly, the V3-V4 regions of the 16S rRNA gene were amplified (15×15 cycles) following a previously described two-step protocol [34] using forward and reverse primers 341F-785R [35]. Purification of amplicons was performed by using the AMPure XP system (Beckmann). Next, sequencing was performed with pooled samples in paired-end modus (PE275) using an MiSeq system (Illumina, Inc.) according to the manufacturer’s instructions and 25 % (v/v) PhiX standard library.

### Microbial profiling

Data analysis was performed as previously described [36]. Processing of raw-reads was performed by using the IMNGS pipeline based on the UPARSE approach [37]. Sequences were demultiplexed, trimmed to the first base with a quality score <3 and then paired. Sequences with less than 300 and more than 600 nucleotides and paired reads with an expected error >3 were excluded from the analysis. Trimming of the remaining reads was done by trimming 5 nucleotides from each end to avoid GC bias and non-random base composition. Operational taxonomic units (OTUs) were clustered at 97% sequence similarity. Taxonomy assignment was performed at 80% confidence level using the RDP classifier [38] and the SILVA ribosomal RNA gene database project [33]. The microbiome was visually inspected for batch effects in PCA; none were found. The resulting OTUs table was normalized using edgeR.

### The glioma dataset

We used a previously published dataset with 53 samples from glioma patients that included the transcriptome, copy number variation, and data from comparative genomic hybridization (CGH). This dataset contained information about age, localization of the tumor, sex and a numerical grading of the severity of the tumor (See Table 1) [31, 39].

**Table 1:**
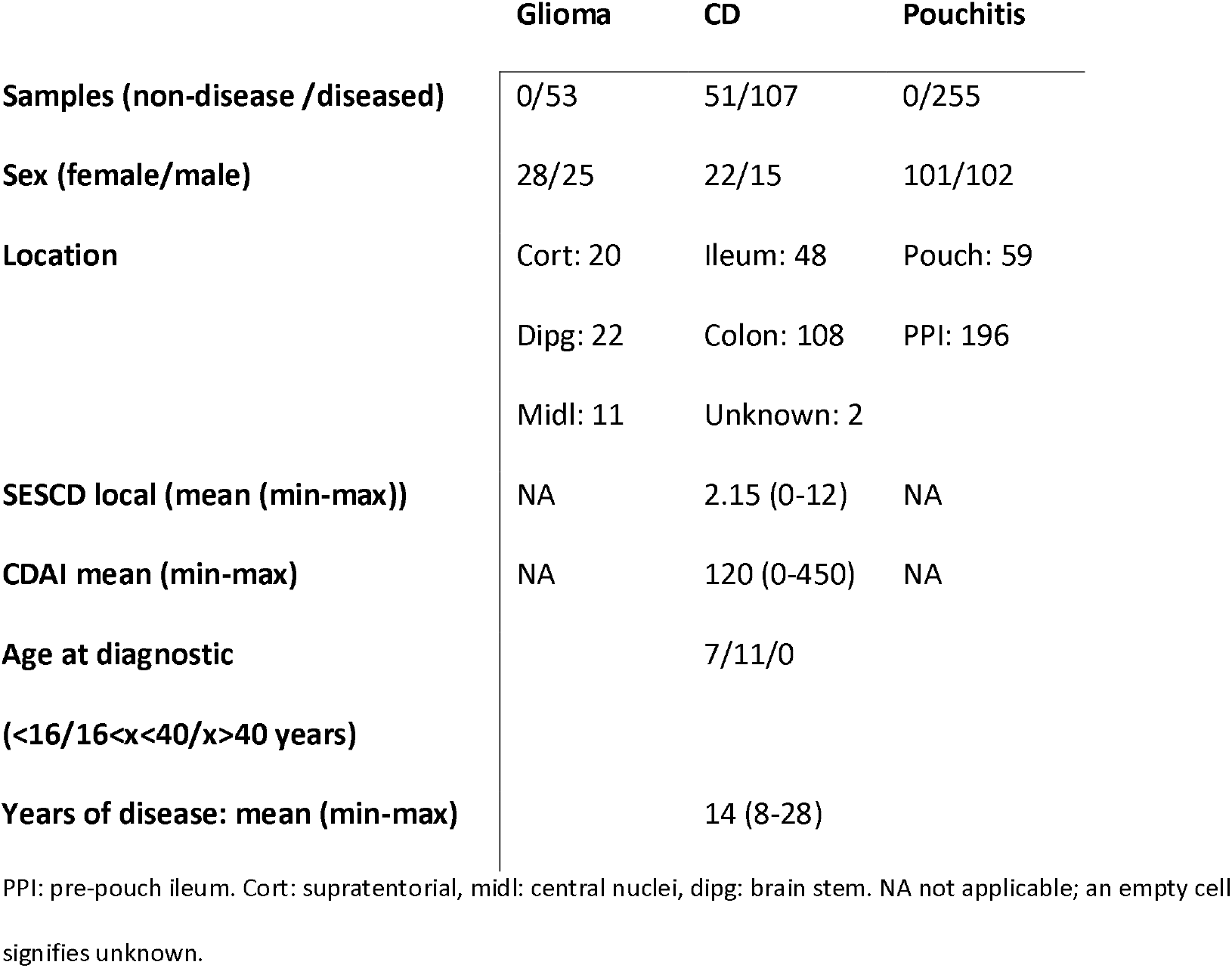
Summary of samples and characteristics of the datasets used.

### The Crohn’s disease dataset

Samples that had both RNA and microbial DNA corresponding to the same patient were included. In total, 158 samples were used for the integration, including those from 18 patients with CD and 19 non-IBD controls (See Table 1). In addition to the samples, clinical information such as age, sex, treatment, years since disease diagnosis, prior surgery, location of the biopsies, segmental simple endoscopic score for Crohn’s disease (SESCD), time of the transplant and response to treatment were collected.

### The pouchitis dataset

A previously published dataset of pouchitis was downloaded containing gene expression data from 273 samples and 16S data on the microbiome presence (See Table 1)[32]. This dataset contained identifiers for the patients, whether the sample was from the pre-pocuh ileum or from the pouch, the sex, the outcome of the procedure and an inflammatory severity score ISCORE. A total of 255 samples from 203 patients were used with data for both transcriptome and microbiome.

### Integration

Sparse regularized generalized canonical correlation analysis (SRGCCA), implemented in RGCCA package (version 2.12), was used for this integration [40]. This variation of the RGCCA method is better suited for biological data with sparsity such as the results obtained by RNA sequencing. The scheme used to add the different canonical components was the centroid scheme, which allows one to determine the positive and negative related variables. The regularization parameters used were those suggested by the tau.estimate, which is a compromise between correlation and covariance [41]. When looking for the covariance from phenotypic categorical variables, one was used for regularization in order to maximize the covariance instead of the correlation.

Numeric values from the same assay were set on the same block. Relevant clinical variables were grouped in one block unless otherwise indicated. Categorical data was encoded as binary (dummy) variables for each factor, where 0 indicates not present and 1 present. Each block was standardized to zero mean and unit variances, and then divided by the square root of the number of variables of the block with the function scale2.

### Hyperparameters testing

The sparse canonical correlation analysis has three hyperparameters: the regularization parameter, tau, the model and the scheme. To evaluate the effect of each hyperparameter, the parameter being tested was changed while keeping constant all the other parameters. This model includes weights indicating the relationship between the blocks.

All models were analyzed using weights from 0 to 1 in the relationship between blocks. To test the effect of the model, all combinations of weights were tested. The inner AVE was used to select the best model. Inner average variance explained (AVE) is defined by how well the components of each block correlate with each other [29].

The scheme controls how the different correlations of the canonical components are summarized. The three schemes available (horst, centroid and factorial) are compared regarding their inner AVE and the selected genes using a simple model.

The regularization parameter, tau, was tested between the minimum accepted value for each block and one for each block of the glioma dataset.

Models were validated using 1000 bootstraps with resampling to assess the stability of the inner and outer AVE. Outer AVE is defined by the correlation between the variables of a block and the component of the block [29].

### Models used

Different models were tested for integration with the same data of the CD and pouchitis dataset. The first model, model 0, used only two blocks, the microbiome and the transcriptome data with interaction between them and with no within interactions (Model not shown).

The second family of models (models 1, 1.1 and 1.2; see Table 2), in addition to the microbiome and transcriptome data, used the clinically relevant variables including some that were related to disease activity. For instance, the CD dataset included the following variables: patient ID, sex, age, age at diagnosis, surgery, treatment, time after transplant and location of the sample.

**Table 2:** Table representing the family of type 1 models.

|  | Transcriptome | Microbiome | Variables |
| --- | --- | --- | --- |
| Transcriptome | 0 | [0-1] | [0-1] |
| Microbiome | [0-1] | 0 | [0-1] |
| Variables | [0-1] | [0-1] | 0 |

[0-1] indicates that the relation between the blocks has been tested at several weights between 0 and 1.

The last family of models (models 2, 2.1, 2.2 and 2.3) used the same information as that for type 1 models, but grouped the clinical variables into three blocks, one for demographics, one for time-related variables and one for variables related to localization of the sample (Table 3). Models 1 to 2.3 were modeled to utilize known, clinically relevant variables with the transcriptome and microbiome data available.

**Table 3:** Table representing the type 2 family of models for the CD dataset.

|  | Transcriptome | Microbiome | Sample | Location | Time |
| --- | --- | --- | --- | --- | --- |
| Transcriptome | 0 | [0-1] | [0-1] | [0-1] | [0-1] |
| Microbiome | [0-1] | 0 | [0-1] | [0-1] | [0-1] |
| Sample | [0-1] | [0-1] | 0 | [0-1] | [0-1] |
| Location | [0-1] | [0-1] | [0-1] | 0 | [0-1] |
| Time | [0-1] | [0-1] | [0-1] | [0-1] | 0 |

The same data of the variables block based on the family model 1 is split into 3 additional blocks. [0-1] indicates that the relation between the blocks has been tested at several weights between 0 and 1.

With the glioma dataset, the microbiome block was replaced by the CGH block. In addition to the previously mentioned models, the glioma dataset was also analyzed considering all the variables from the different blocks as a single entity, which is known as a superblock [42].

Only the models in which all the blocks were part of a single connected network were analyzed. For models 1 to 2.3, all the combinations of different weights on the model matrix were analyzed. First weights 0, 0.5 and 1 were used to select the model with the highest inner AVE. To further accurately describe the interactions of models 1.1 and 2.1, different weights from 0 to 1 by 0.1 were tested; the best one resulted from model 1.2 and 2.2, respectively. The higher the AVE is, the better the model is. A direct interaction between the microbiome and the transcriptome was used to check if the results of model 2.2 had improved in model 2.3.

## Results

The effects of the hyperparameters on SRGCCA were first evaluated on the glioma dataset (glioma dataset, Table 1) [31]. Once we had determined the effect of each hyperparameter, we integrated the two different datasets. First, we looked for the relationship between the transcriptome and the microbiome in biopsy samples from patients with CD undergoing HSCT (CD dataset, Table 1). Second, we studied the microbial relationships with the transcriptome in a previously published pouchitis cohort (pouchitis dataset, Table 1) [32]. Then we compared the models on each dataset for robustness.

### Hyperparameters on the glioma dataset

We first analyzed the best strategy to find the right values of the hyperparameters on SRGCCA on the glioma dataset. By hyperparameters we mean the scheme used, the regularization effect, and the models as constructed by weights, all of which can affect the final solution of the SRGCCA.

The regularization parameter, also called tau, controls the number of variables selected by each block. Tau can be estimated by using Schäfer’s method [41], which tries to conserve both the correlation and the covariance. When estimated by this method, the tau provides a good intermediate solution for numeric variables. For those blocks that encode categorical variables as numeric values, it is more relevant the covariance; thus, a tau value of 1 was used. The effect of tau on the inner AVE is shown in Fig. 1.

**Fig. 1.**
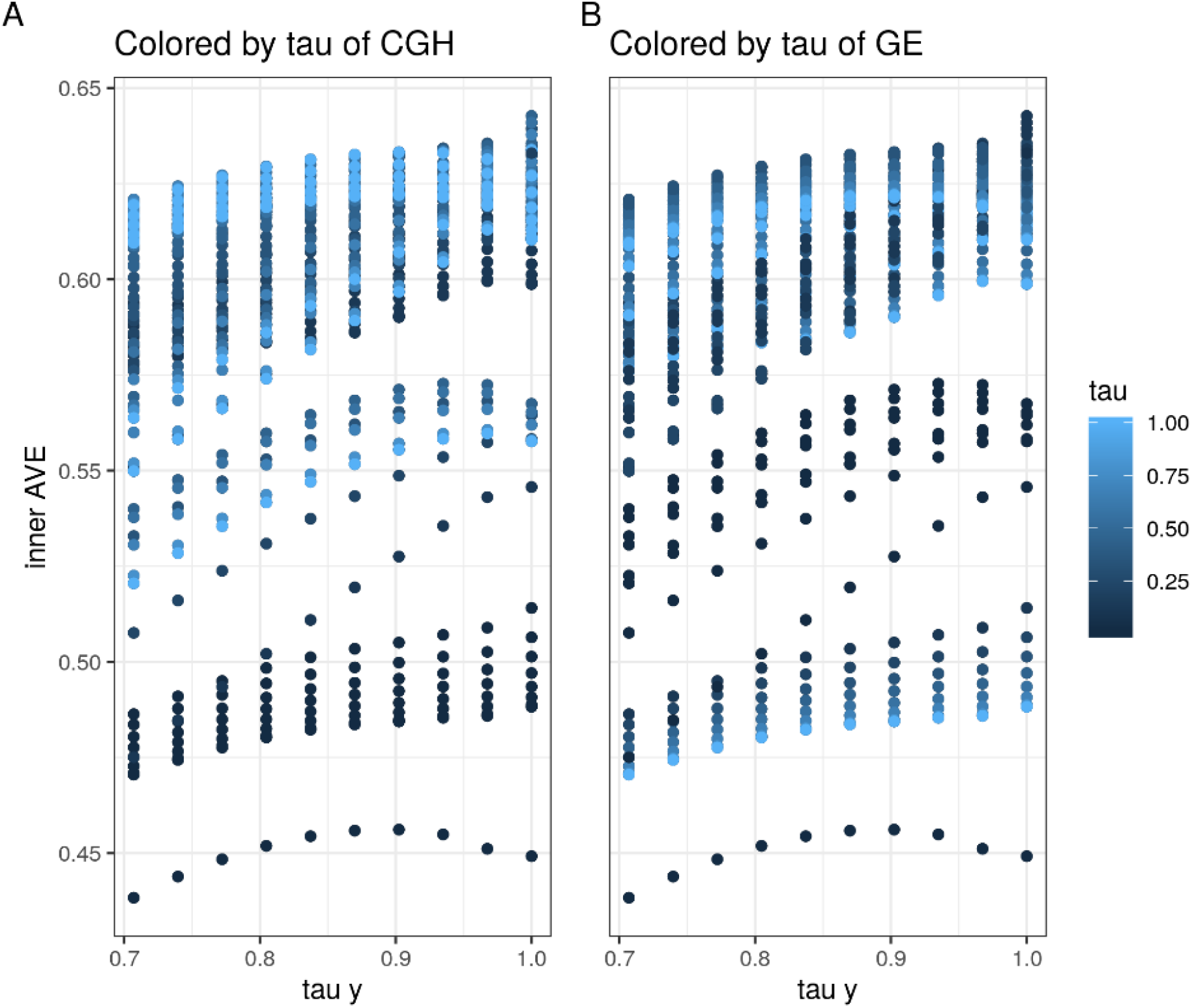
Regularization effect of the inner AVE for the same model on the glioma dataset. Each point is the result of an SRGCCA with different tau values for each block (GE as the transcriptome, CGH (comparative genomic hybridization) for the copy number variation and y for the location).

All of the weights between 0 and 1 (by 0.1) in the glioma dataset were analyzed using all three schemes: horst, centroid and factorial. The horst and the centroid scheme were similar while the factorial resulted in the most different AVE values (see Additional file 1). The centroid scheme was selected because it took into account all the relationship regardless of the sign of the canonical correlation, and because of its similarity to horst.

The three blocks with the best tau and the centroid scheme were analyzed by tuning the weights. According to the inner AVE, the best model was that in which the weights between the transcriptome and location, the transcriptome and the CGH, and the CGH block were linked to variables related to the location with weights of 1, 0.1 and 0.1, respectively.

When we added a superblock to the data, there was an increase of 0.01 on the inner AVE of the model. The model with the superblock that explained most of the variance was that in which the weights of the interaction within the transcriptome, between the superblock and the CGH, between the transcriptome and the localization, and between CGH and transcriptome were 1, 1, 1 and 1/3, respectively. Fig. 2 shows the first two components of the superblock.

**Fig. 2.**
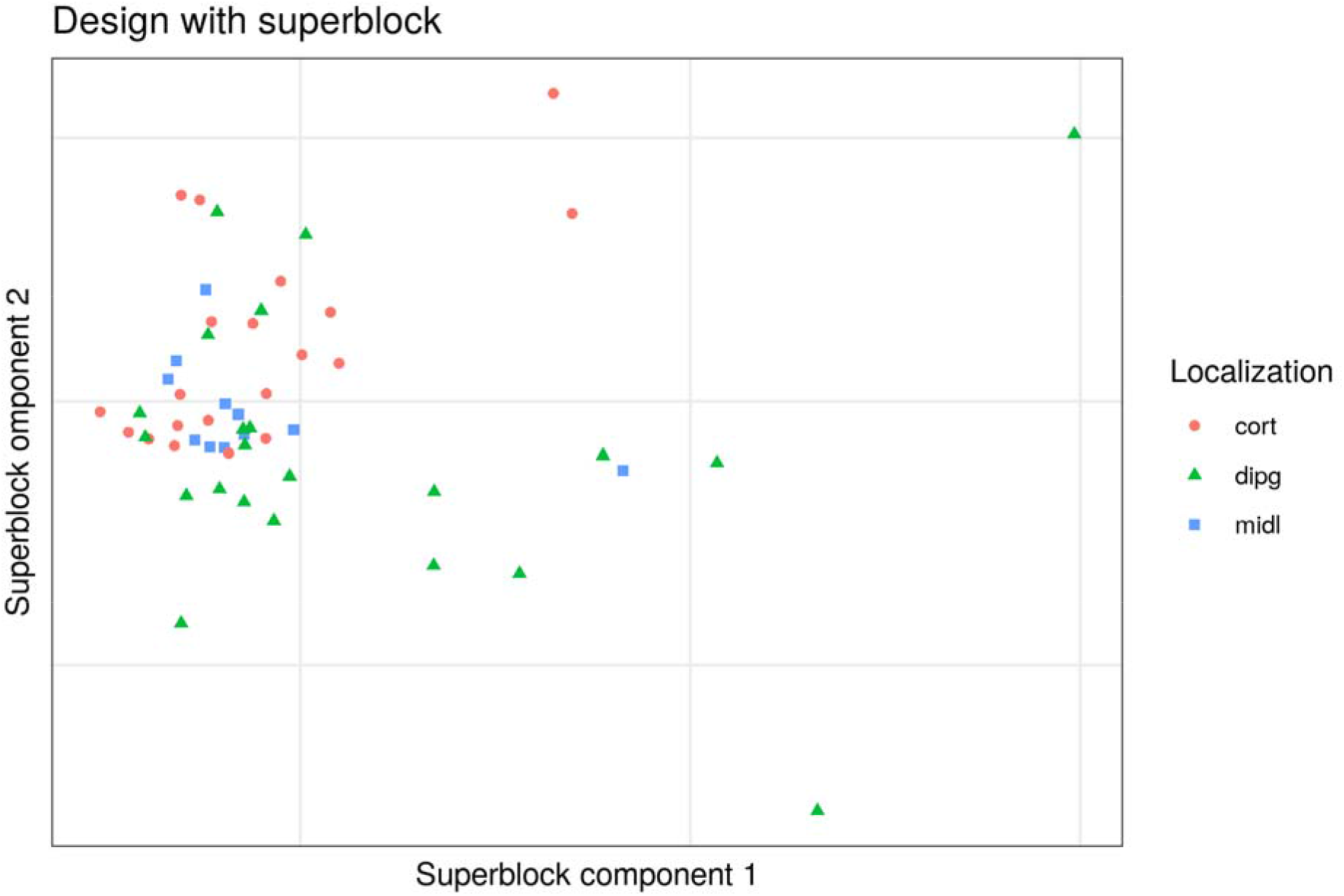
First two dimensions of the superblock on the glioma dataset. The first two components of the superblock with the best model, according to the inner AVE from the glioma dataset. Each point is a sample (colored by location) signifying the following Cort: supratentorial, dipg: brain stem, midl: central nuclei. The locations of the samples could not be determined by the position of the samples on these components

Adding to the model one block containing the age of the patient and the severity of the tumor decreased the inner AVE. The best model with these blocks, according to the inner AVE, was that in which the interactions within transcriptome, between the transcriptome and the localization, between the transcriptome and the CGH and between the CGH and the other variables were 1, 1, 1/3 and 1/3, respectively (See additional file 2, Glioma’s sheet). The first components of each model can be seen in Fig. 3:

**Fig. 3.**
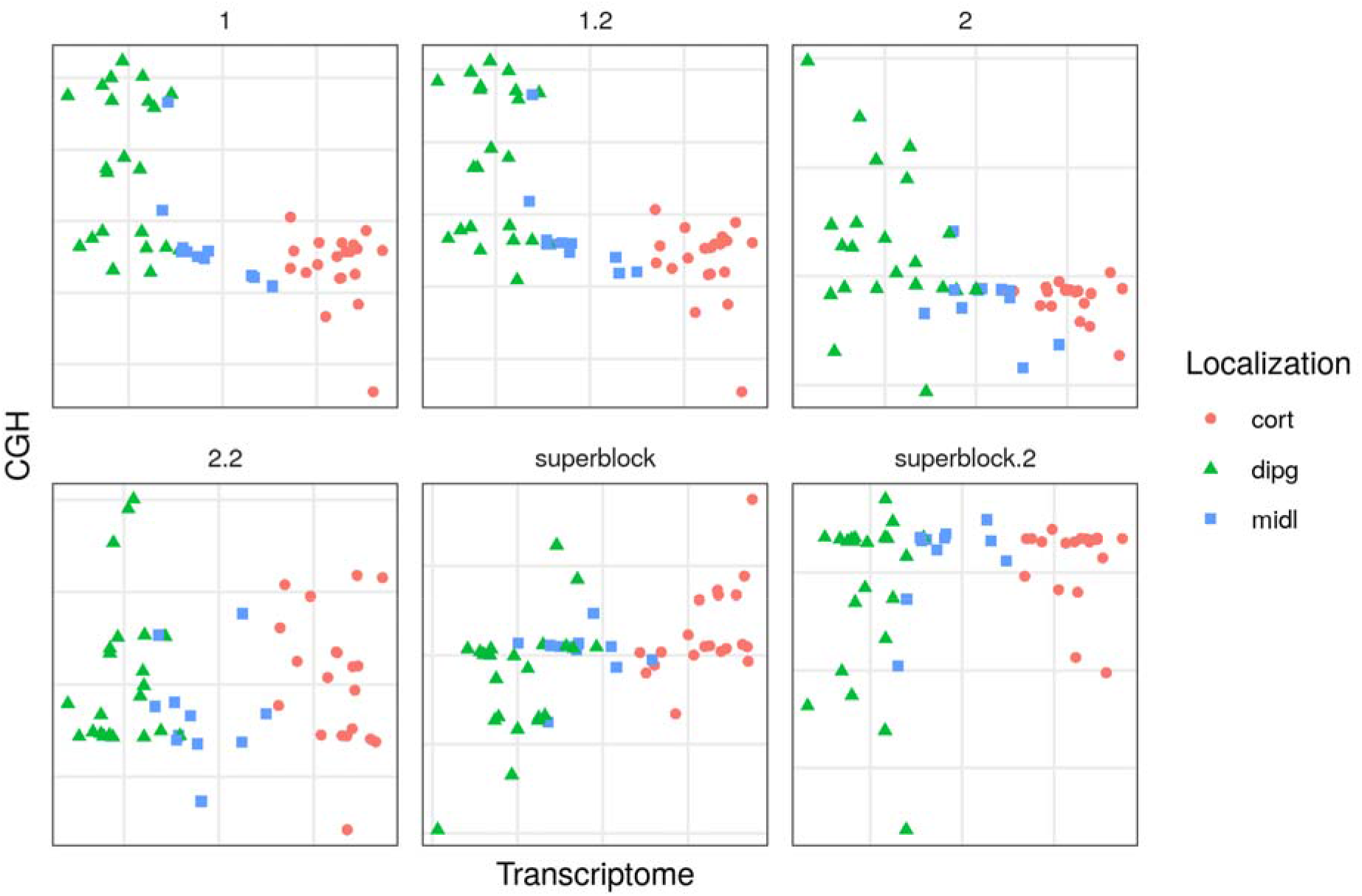
First dimensions of the transcriptome and the CGH block of models on the glioma dataset. Comparison of the different models by visualizing the first components of the transcriptome (GE) and the copy number variation (CGH) blocks from the glioma dataset. Each point represents a sample (colored by location). Cort: supratentorial, dipg: brain stem, midl: central nuclei.

Here, one can see that the tau value proposed by Schäfer’s method is a good approximation of the optimum value for the data and can be used in the other datasets containing omics data. As the model with a superblock did not help explain the relationships between blocks, it could not be used in the other datasets. The scheme selected was the centroid, which takes the absolute value of the relation between components. These hyperparameters were used for further analysis on the CD and pouchitis datasets.

### Analyzing the models on the CD and pouchitis datasets

Each dataset has different relationships that should be taken into account when looking for relationship on the dataset. The CD dataset (see Table 1) was analyzed with SRGCCA. The samples are shown on the first components of the microbiome and the transcriptome **Error! Reference source not found.Fig. 4**). The first canonical components for the microbiome and the expression data for all of the models facilitated the classification of the samples according to the sample location (ileum or colon) or disease status (CD or control).

**Fig. 4.**
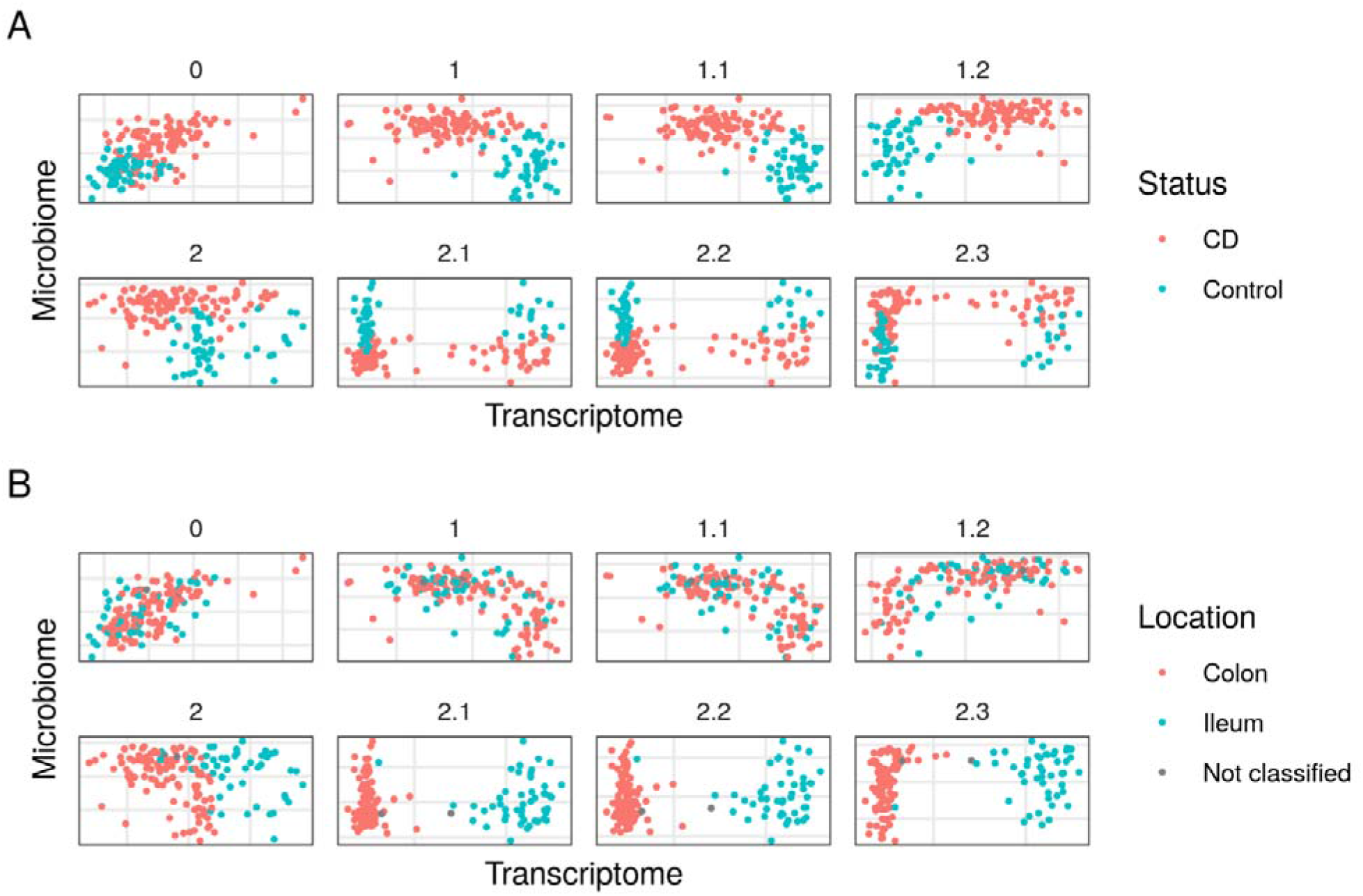
First dimensions of the transcriptome and the microbiome block of models on the CD dataset. Comparison of the models that better explained the interaction between the microbiome and the transcriptome data on the CD dataset. Each point represents a sample (colored by disease status): A, non-CD (Control) or CD; and B, by location, colon or ileum, on the first components of the transcriptome and the microbiome.

Model 1.2 had the highest inner AVE of the family 1 model. A search for the highest inner AVE within the family 2 models resulted in model 2.2, which revealed a direct relationship between the transcriptome and the location-related variables, while the microbiome was associated with the demographic and location-related variables.

The pouchitis dataset (See Table 1) was analyzed and the samples are shown on the first components of the microbiome and the transcriptome in Model 1.2 had the highest inner AVE. A search for the highest inner AVE among the family 2 models resulted in model 2.2, although it does not have the highest inner AVE. Moreover, no direct relationship between the transcriptome and the clinically relevant variables was apparent (See additional file 3). Family 2 models better stratified the samples by location than did those of family 1.

In the CD dataset we see that the relationships evident in the model affected the distribution of samples on the components of both the transcriptome and the microbiome. We found that model 2.2 best stratified the disease status and locations of the samples. Other models grouped the samples by disease status and location based on how close the model was to the weights associated with model 2.2. In all of the models we observed associations between the disease status and the microbiome and the sample location with the transcriptome.

With the pouchitis dataset no stratifications by sex were similarly apparent to that observed in the CD dataset. Nonetheless, they were separated by location-related variables in some models, albeit not as clearly as with the CD dataset. This might indicate that while sex does not affect the interaction, the location-related variables do affect the pouchitis.

### Comparison of models in Crohn’s disease and the pouchitis dataset

The same dataset with different models results in different relevant variables. With the CD dataset we looked for the best model using a single block for the clinically relevant variables, following the family model 1 structure. The family 1 model with the highest AVE was that in which the transcriptomics was related the phenotype by 0.1, while the microbiome was related to the clinically relevant variables by 1. This model revealed that the relationship between the microbiome and the clinically relevant variables carried more weight than that between the clinically relevant variables and the transcriptomics on the CD dataset.

The best model according to the inner AVE score on the CD dataset was model 2.2. In this model, the transcriptome was related to location-related variables by a weight of 1, while the microbiome was related to demographic variables, and to location related variables, by a weight of 1 and 0.5, respectively. Demographic variables were also linked by 1 to the time variables block (See additional file 2, CD’s sheet).

The interaction of genes within the transcriptome was also analyzed on the CD dataset. It increased the inner AVE score between 0.10 and 0.03 depending on the model. However, it was not deemed important to find the relationships between the transcriptome and the microbiome and thus was not compared between datasets.

In order to analyze the accuracy of the models, one thousand bootstraps were used to integrate the data from the CD dataset (see Fig. 6 and Table 4 below). Model 2.2 had both higher inner and outer AVE mean values and less standard deviation (Fig. 6 and Table 4). This indicates that it was more robust than the other models, regardless of the input data.

**Table 4:** Bootstrapped mean and standard deviation of inner and outer AVE values on the CD dataset.

| Model | AVE | Mean | Sd |
| --- | --- | --- | --- |
| 0 | inner | 0,550 | 0,0469 |
| 1.2 | inner | 0,768 | 0,0223 |
| **2.2** | **inner** | **0,785** | **0,0163** |
| 0 | outer | 0,104 | 0,0132 |
| 1.2 | outer | 0,088 | 0,0106 |
| **2.2** | **outer** | **0,105** | **0,0069** |

The best models according to the mean are shown in bold.

On the other hand, the model with the highest inner AVE on the pouchitis dataset was model 1.2, which included a relationship between the microbial data and the transcriptome of 0.1, as well as a relationship between the microbial data and the metadata of 1 (See additional file 2, Pouchitis’ sheet). Other models from family 2 had less AVE. The family 2 model with a higher inner AVE was model 2.2. The first dimension of the transcriptome separates by location on models of family 2 while the first component of the microbiome doesn’t separate by males and females (See Fig. 5).

**Fig. 5.**
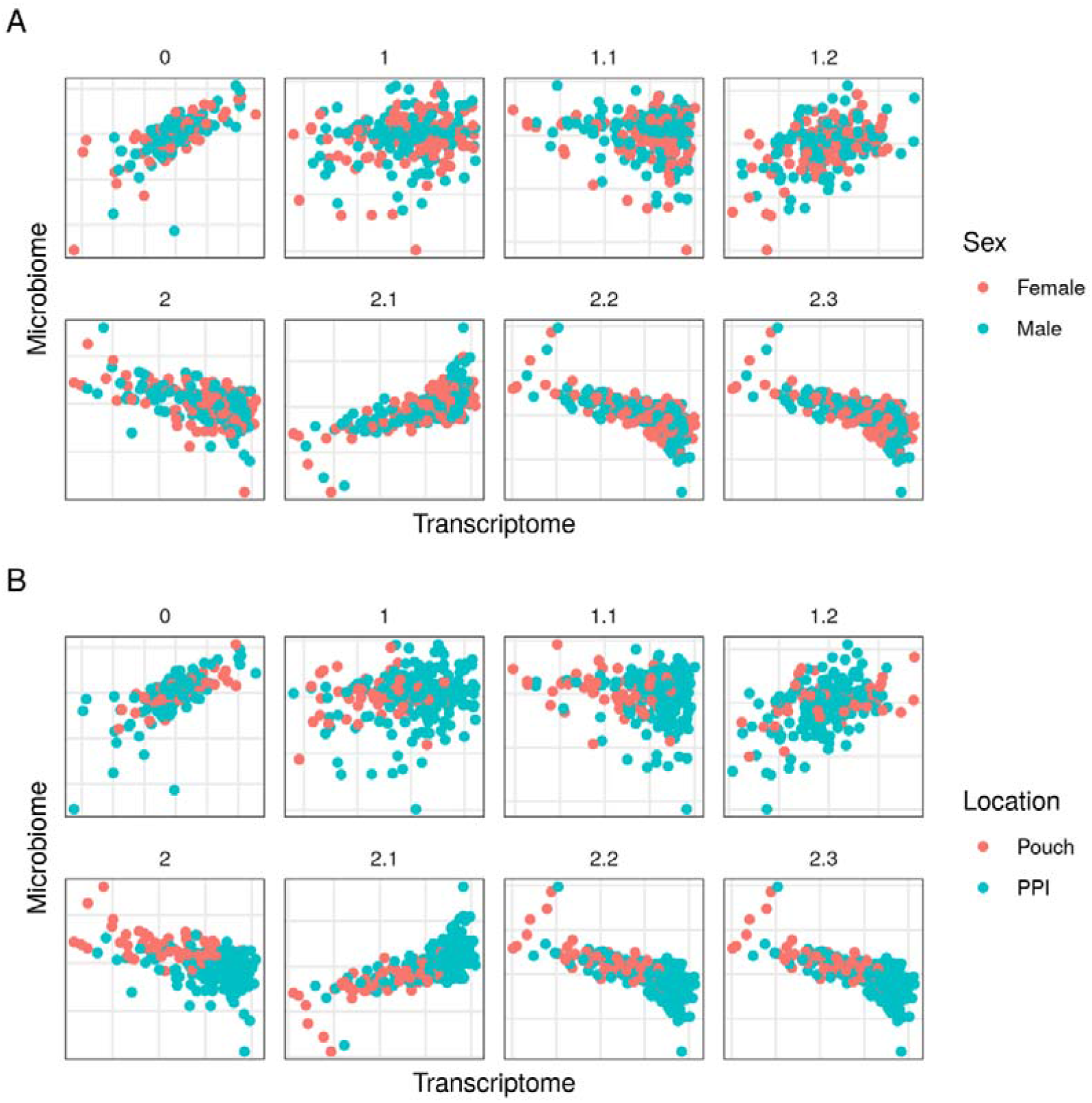
First dimensions of the transcriptome and the microbiome block of models on the pouchitis dataset. Comparison of the models vis-a-vison the pouchitis dataset by the first component of the transcriptome and the microbiome from the CD dataset. Each point represents a sample colored by sex (A), where females are in red and males in blue, and by location (B), where the pouch is the red, and PPI is the pre-pouch ileum

The bootstrap analysis of the one thousand bootstraps on the pouchitis dataset showed that model 1.2 had the highest mean inner AVE, although model 0 had the highest mean outer AVE (See Table 5). As the mean outer AVE for model 1.2 was close to model 0, the former was considered the most robust.

**Table 5.**
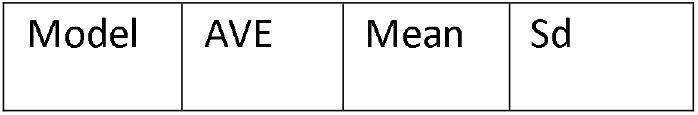
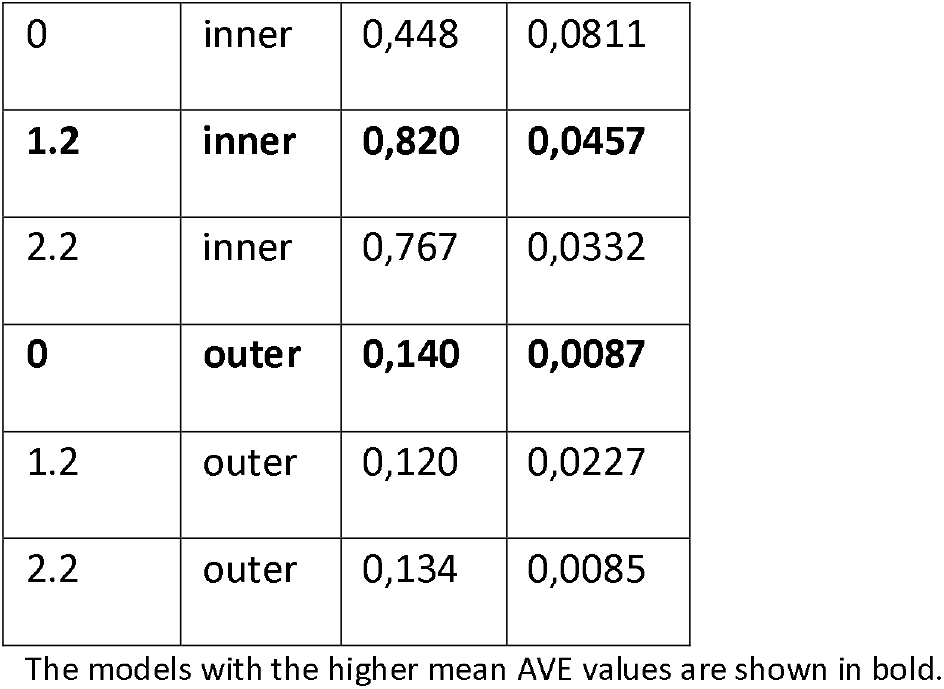
Bootstrapped mean and standard deviation of inner and outer AVE values on the pouchitis dataset.

| Model | AVE | Mean | Sd |
| --- | --- | --- | --- |
| 0 | inner | 0,448 | 0,0811 |
| **1.2** | **inner** | **0,820** | **0,0457** |
| 2.2 | inner | 0,767 | 0,0332 |
| **0** | **outer** | **0,140** | **0,0087** |
| 1.2 | outer | 0,120 | 0,0227 |
| 2.2 | outer | 0,134 | 0,0085 |

The models with the higher mean AVE values are shown in bold.

The models with the highest inner AVE were more robust to different data, which indicates that they can be applied more generally and not solely to these samples.

## Discussion

This study provides a framework for identifying interactions between blocks of data, a step towards understanding biological relationships between datasets or between datasets and other particularly relevant variables. First, we studied the hyperparameters’ influence on a glioma dataset, adjusting their values. Then, we developed a method to find the best model for the relationships between blocks. Lastly, we validated the method in two independent cohorts.

We explored the regularization of the blocks on the glioma cohort. The regularization of a block modulates how many variables are selected [26, 28]. The use of tau 1 allowed us to select all variables, which maximized the covariance of the variables. On blocks that included only clinically relevant categorical variables, regularization must be equal to 1. As the transcriptome and microbiome blocks contain many variables, a shrinkage parameter close to 0 was expected, as was observed with the glioma and other cohorts. In addition, estimating tau for the quantitative blocks resulted in higher inner AVE scores since the quantitative variables that contributed most to the data variation were selected.

Based on the regularization obtained, we explored different schemes of integration on the glioma cohort. The resulting canonical components of the centroid and horst schemes did differ in some models. In fact, the canonical correlations between blocks was likely positive, making the differences between these two schemes unobservable. The centroid scheme was selected to analyze the CD and the pouchitis datasets, since canonical correlations are not always positive.

Independently of the scheme involved, a superblock not only aids in interpretation, but also helps account for the possibility of interactions between variables of the same block. The increase observed in the inner AVE may have stemmed from the interaction between variables of the same block. However, such an interpretation is not as clear as with blocks generated by a single assay or from closely related variables [28]. The superblock, which is used for redundancy analysis, did not help in terms of grouping different samples [42]. Moreover, if the goal of the model is to accurately represent the system under study, the superblock is not necessary, regardless of the assistance it provides in improving the inner AVE.

The superblock is usually related to all the other blocks. Typically, a weight of 1 is used to indicate a direct relationship between two blocks. Modifying the weights of the model influenced the result by changing AVE scores and the variables selected from each block. The highest inner AVE score was not defined by the highest weights on all the relationships.

The weights of the models represent how much one block interacts with another if the interactions are linear, an assumption of any canonical correlation [29]. In such cases, the weights are representative of the interactions between blocks.

The weights define the relationships between blocks in SRGCCA, which together determine the model of the components. Other methods like MCIA and JIVE assume a common relationship between all components, which results in a common space for the samples [25, 26]. This difference is crucial for exploring the role of the components; for example, in this paper each model represents the same system with different interactions and assumptions. Comparing different models after the SRGCCA led to explanations for different aspects of the same system.

Looking at the glioma data, the best model according to the inner AVE was that with the superblock. As previously explained, this model might represent the hierarchical relationships present in the data. However, the superblock did not provide more interpretable results in the glioma dataset.

In the glioma cohort, the model lacking the superblock but with the highest inner AVE indicated that the localization of a tumor influences the transcriptome to a greater degree than the copy number variations, if the relationships are linear. Adding additional information on the samples’ localization origin did not increase the inner AVE, suggesting that there was a high dependence between localization and the tumor transcriptome.

Interactions within the transcriptome usually increase the inner AVE of the models. With the CD and the pouchitis datasets, self-interaction increased the inner AVE, as well as the selected features, except in models 0 to 1.2 in the CD data set. This suggests that the interactions within the same omic block become relevant if the model does not take into account the interaction between other clinically relevant variables. If other relevant variables are included, then the effect of this interaction is significantly less.

Model 0 looked for direct relationships between the microbiome and the transcriptome. Confounders that influence both transcriptome and microbiome, such as age or the localization and inflammation status, were not taken into account in this model. This is due to the fact that they can bias the relations found with this model [43]. Nonetheless, this model was capable of grouping the samples of the CD cohort according to their disease status, though this was not true of the pouchitis dataset.

Family 1 models use three blocks, including one for clinically important information about the samples. This new block was added to avoid biasing the integration by known factors of the samples. In the best model of this family, the microbiome block had a weak relationship with the transcriptome. This weak relationship was possibly indicative that the relations were not lineal. If the relationships were not lineal, then they could not be fully identified by RGCCA [29]. Another possibility is that the microbiome was related to other variables not included on the dataset.

Finally, family 2 models, compared to those of family 1, were designed to explain the relationship between the microbiome and transcriptome, allowing for the presence of independent interactions with location, age and other demographic-related variables. In family 1 models all the relevant variables were mixed together. In order to allow for such interactions, unrelated variables were separated in different blocks. Genes selected by SRGCCA proved different between the family 1 and 2 models (see Fig. 7), suggesting that the relationship between microorganisms and genes was heavily influenced by location, time and demographic-related variables (see Fig. 8).

**Fig. 6.**
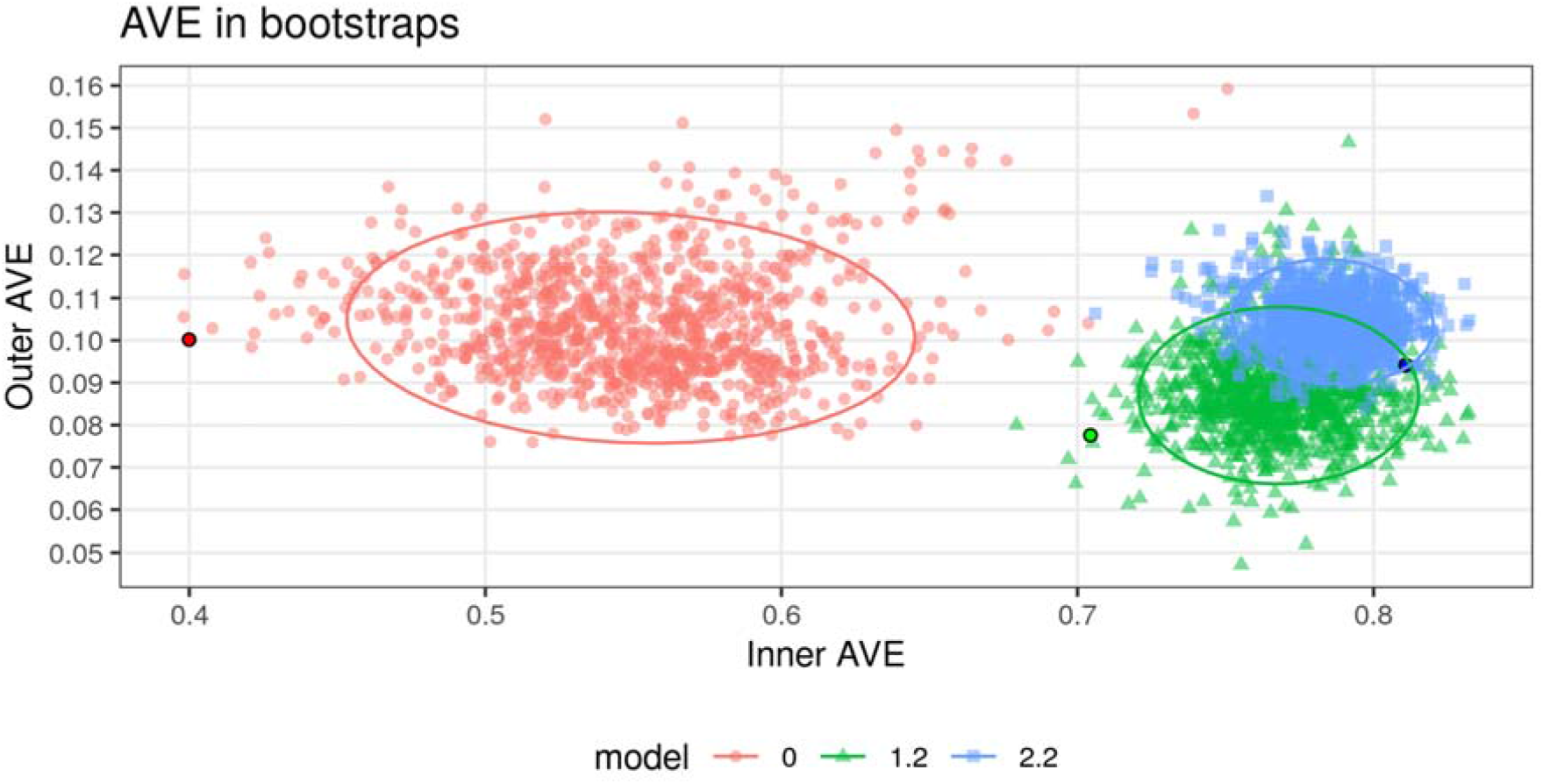
Bootstrap results of three models on the CD dataset. Variance of AVE using the same samples on three models with the CD dataset. Each point shows the AVE for each analysis performed. The brighter colors reflect the result of this model on the original data (including all samples).

**Fig. 7.**
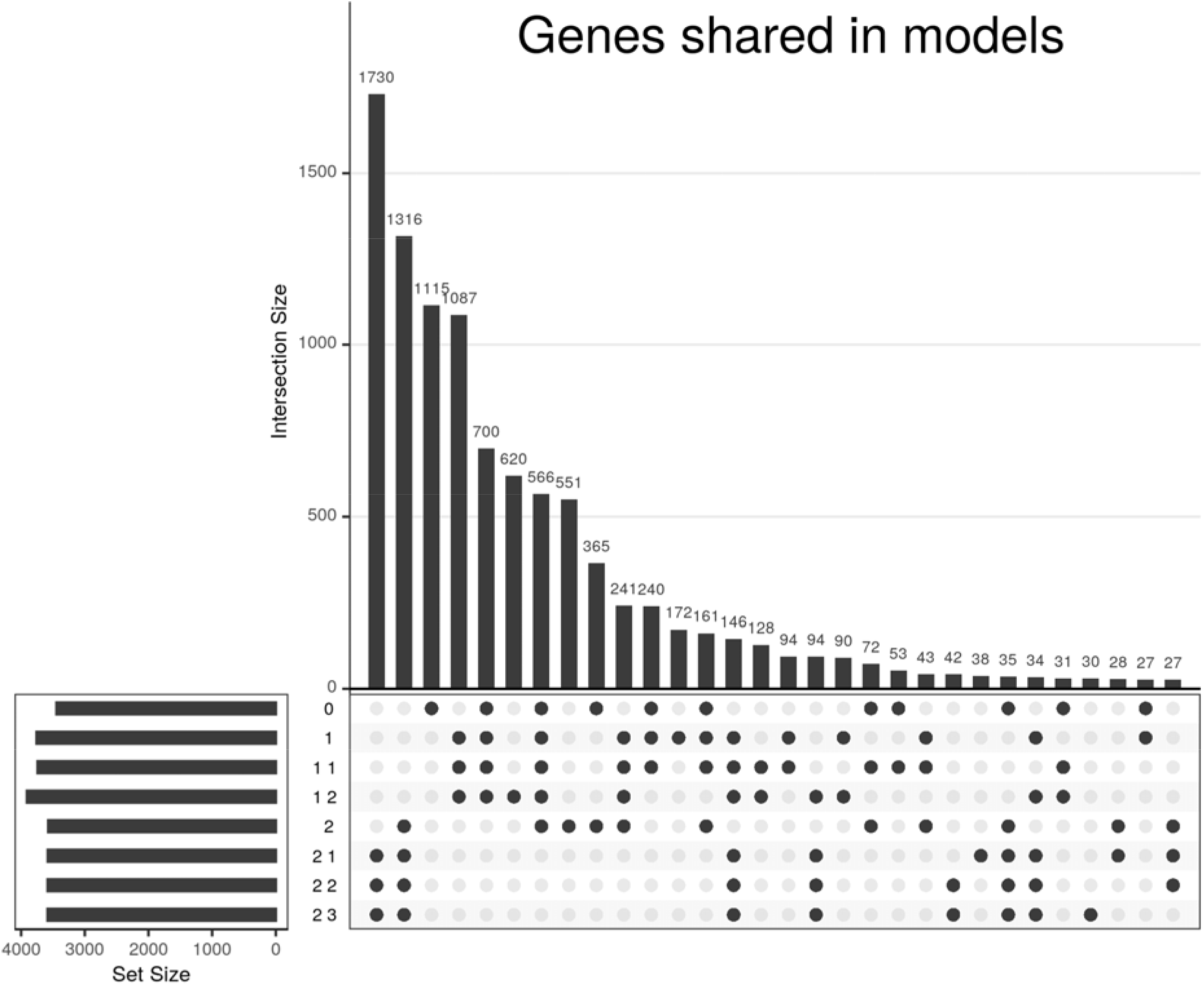
UpSet plot of the genes shared between models on the CD dataset. The heights of the bars represent the genes shared between the models selected by the points; 30 intersections are shown. The lengths of the horizontal bars represent the selected genes in each model.

**Fig. 8.**
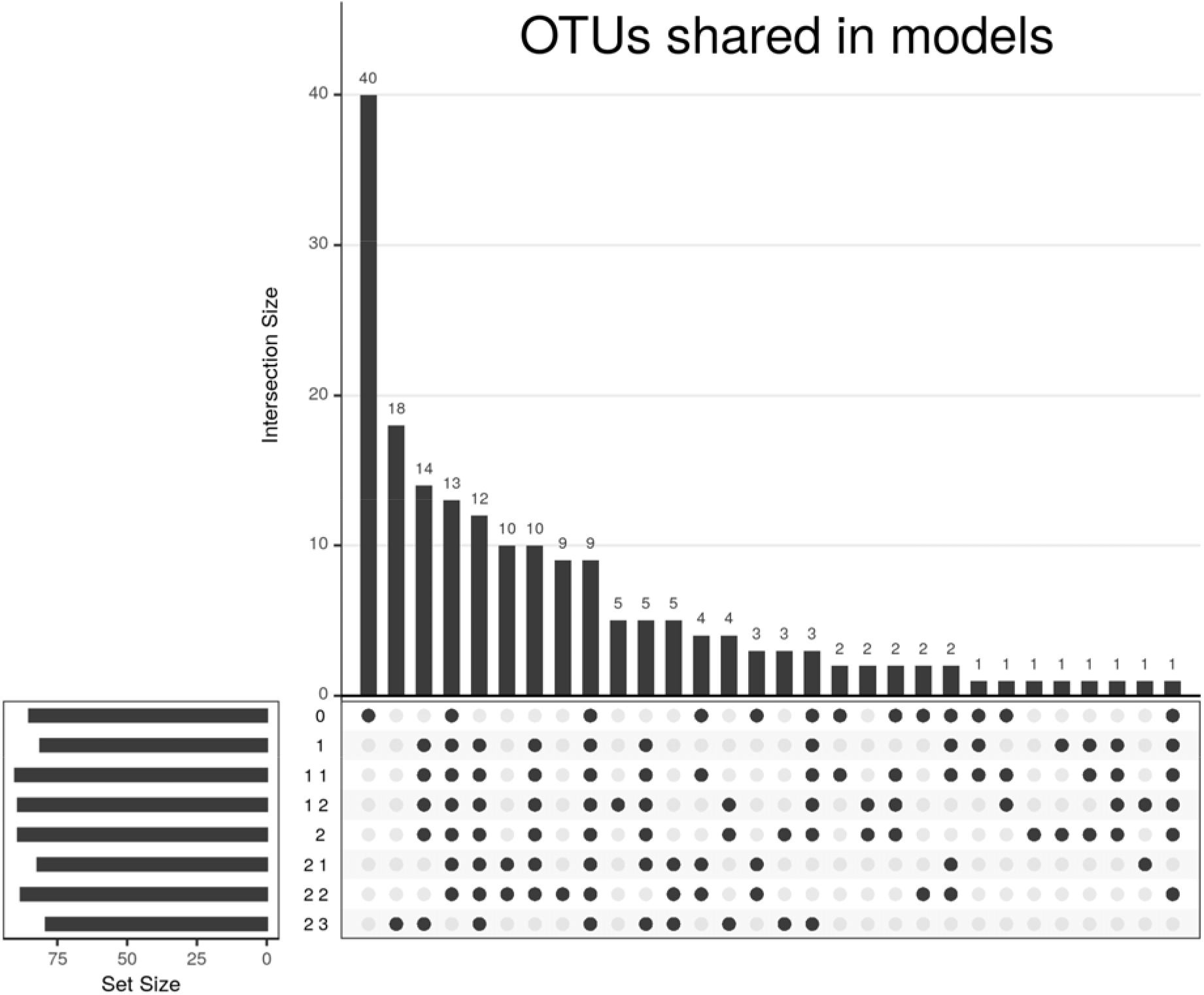
UpSet plot of the microorganisms selected by the models on the CD dataset. The heights of the bars represent the OTUs shared between the models selected by the points; 30 intersections are shown. The lengths of the horizontal bars represent the selected OTUs in each model.

Of all these models, the best according to the inner AVE on the CD dataset was model 2.2. This model showed known differences between the transcriptome in the gut regions [15]. The microbiome separated the samples by disease status, indicating that it was highly relevant for the relationship with the transcriptome. In addition, the dispersion of mean average variance was reduced as more complex models were used, as can be seen in Fig. 6, indicating that they were more robust to different data.

In the CD dataset, a cursory analysis confirmed that the genes selected by SRGCCA with model 2.2 were related to the sample location [15]. Among the selected microorganisms previously linked to CD dysbiosis on this list were Faecalibacterium sp. and Bacteroides sp. (see Additional file 4)[44]. This suggests that the variables selected were relevant for their role in both the tissue and the disease. Thus, the genes and microorganisms that have significant relationships, in this context, were likely to be present.

In the pouchitis dataset, model 1.2 captured a greater degree of variance than model 2.2, contrary to the results obtained with the CD dataset. This might be because potentially important variables, such as age, were lacking and possibly because the model was confounded. In addition, we could not make direct comparisons with the CD dataset as it did not include non-diseased samples. This is due to the fact that the model differentiates by subgroups of patients instead of by a distinct relationship between healthy and diseased samples.

The findings of this study have to be assessed in light of certain limitations. RGCCA can not describe a causal relationship or the mechanisms underlying the relationships between RNA transcriptomics and the microbiome. However, models for RGCCA can be used to select variables for further studies and experiments in order to validate these relationships.

When examining an interaction within a block, we only assumed the existence of an interaction within the transcriptome. However, it must be noted that microorganisms create communities for which the interactions of several microorganisms is essential and we did not consider interaction within the microbiome in the present study [45]. Knowing how microbial communities rise and interact remains an open question that could affect any interpretation of the results [45, 46].

In the present study, as we did not use a simulated data set with known relationships between blocks, we could not asses the specificity or sensitivity of our approach. In addition, we did not confirm by further analysis and experiments whether the selected variables were necessary to start or maintain CD or pouchitis.

## Conclusions

RGCCA is a powerful integration tool. We have shown that the model is the most important parameter when selecting variables. The weights of the model represent the strengths of the relationships between blocks. Here we propose a robust methodology to identify the best models guided by the inner AVE when there is no prior knowledge of the existing relationship.

This method can identify relationships in complex systems such as Crohn’s disease by taking into account the interactions between the microbiome, transcriptome and the relevant clinical variables. The resulting analysis can improve our understanding of the biological relationships between different omics datasets and other relevant (clinical) variables.

## Data Availability

Data available: glioma data at https://biodev.cea.fr/sgcca/.The datasets supporting the conclusions of this article are available in the Gene Expression Omnibus repository, https://www.ncbi.nlm.nih.gov/geo/query/acc.cgi?acc=GSE139179 (RNA-seq) and https://www.ncbi.nlm.nih.gov/geo/query/acc.cgi?acc=GSE139680 (microbiome) for
the CD dataset and https://www.ncbi.nlm.nih.gov/geo/query/acc.cgi?acc=GSE65270 for the pouchitis dataset and its additional file(s).

https://biodev.cea.fr/sgcca/

https://www.ncbi.nlm.nih.gov/geo/query/acc.cgi?acc=GSE139179

https://www.ncbi.nlm.nih.gov/geo/query/acc.cgi?acc=GSE139680

https://www.ncbi.nlm.nih.gov/geo/query/acc.cgi?acc=GSE65270

## Abbreviations

IBD: inflammatory bowel disease
CD: Crohn’s disease
RGCCA: regularized generalized canonical correlation analysis
SRGCCA: sparse regularized generalized canonical correlation analysis
AVE: Average variance explained
CGH: Comparative genomic hybridization
HSCT: hematopoietic stem cell transplantation
SESCD: simple endoscopic score for Crohn’s disease

## Declarations

### Ethics approval and consent to participate

The protocol was approved by the Catalan Transplantation Organization and by the Institutional Ethics Committee of the Hospital Clinic de Barcelona. All patients provided written consent following extensive counselling.

### Consent for publication

Not applicable.

### Availability of data and materials

Analysis code available at: https://github.com/llrs/TRIM repository.

Parameter studies performed at https://github.com/llrs/sgcca_hyperparameters.

Helper package required for the analysis available at https://github.com/llrs/integration-helper. Package with the methodology:

Project name: inteRmodel

Project home page: https://llrs.github.io/inteRmodel/

Operating system: Platform independent

Programming language: R

License: MIT

Data available: glioma data at https://biodev.cea.fr/sgcca/.

The datasets supporting the conclusions of this article are available in the Gene Expression

Omnibus repository, https://www.ncbi.nlm.nih.gov/geo/query/acc.cgi?acc=GSE139179 (RNA-seq) and https://www.ncbi.nlm.nih.gov/geo/query/acc.cgi?acc=GSE139680 (microbiome) for the CD dataset and https://www.ncbi.nlm.nih.gov/geo/query/acc.cgi?acc=GSE65270 for the pouchitis dataset and its additional file(s).

### Author contributions

LR analyzed the data and wrote the first draft of the manuscript. AiM performed the DNA and RNA extraction and participated in the microbiome analysis. AMC performed the RNA mapping and cleaning. JJL provided guidance and technical assistance. ET, AC and ME recruited the non-IBD patients for the study. JP, ER, recruited IBD patients for the study. AS selected the clinically relevant variables, provided guidance on model selection and data interpretation and corrected the manuscript. MC contributed to samples processing. DH and AmM performed the microbiome sequencing. All authors read and approved the submitted manuscript.

## Acknowledgements

We thank Daniel Aguilar for RNA-seq analysis assistance and Ilias Lagkouvardos for his assistance on 16S-seq analysis. We are grateful to Joe Moore for English-language assistance.

